# The Electronic Health Record System May Destroy The Empathy

**DOI:** 10.1101/2023.04.07.23288307

**Authors:** Mohammadreza Nilipour Tabatabaei, Seyed Amirhossein Dormiani Tabatabaei

**Affiliations:** Islamic azad university,Najafabad branch, Isfahan,Iran; Isfahan University of Medical Sciences isfahan,Iran

**Keywords:** Empathy, Patients, Physicians, Computer, Communication, Hematology, Neoplasms

## Abstract

**Introduction:** The rapid growth of the Electronic Health Record (HER) systems has affected our understanding of the EHR while still providing compassionate health care and optimizing patient-physician communication. Empathy as a core component of this communication has been connected to other interpersonal interaction indicators such as trust and patient satisfaction.The vulnerable situation of patients with hematologic malignancies necessitates effective empathetic interaction with full attention from the physicians and those working in oncology wards.

**Methods and materials:** Patients were enlisted from the Hematology-oncology ward and Clinic. (either new referrals or follow-ups).

120 patients were stratified into two arms of the study asking them to observe short videos and complete the questionnaire regarding the physicians: one uses an Electronic Health Record system and another consults the patient without an Exam Room Computer. patients were asked to state the level of their agreement or disagreement with each of the statements of the Persian translation of the Jefferson Scale of Patient Perceptions of Physician Empathy (JSPPPE) questionnaire.

**Results:** Patients viewed the EHR(#1) and No Computer (#2) videos for a crossed-over clinical trial. The No Computer visit resulted in significantly better empathy scores compared with the EHR visit.

**Conclusion:** Based on the results of this study, The Empathy phenomenon at its core will never change (48,49)but has various facets that are progressively being understood. we continue to advance technological devices to improve the foundation of patient care and outcomes. If medical care trends in The triumphs of technology, especially in hematooncological clinics, continue as expected, empathy will become an even more critical issue.

## Introduction

the electronic health record (EHR) is becoming more widespread in face-to-face health encounters and is using in clinical settings for managing and keeping patient data and billing processes(1,2).Iran’s health ministry has announced that the implementation of the EHR officially started in February, 2017 and applied through 235 cities in November 2018, in addition, the spread of Covid-19 has accelerated it more. In January, 2021, the progress of electronic health records was about 75%, which reached over 90% due to the legal requirement. according to experts, the EHR has reduced care costs, prescription mistakes, and medication errors and ultimately improves drug therapy and patient health(3,4,5). this rapid growth has affected our understanding of the EHR.while still providing compassionate health care and optimizing patient-physician communication plays a principal role in each phase of every teamwork; fundamental factors that influence clinician-patient communication as teamwork in this clinical encounter,should be Scrutinized in more depth than previously several studies(6).

Empathy as a core component of this communication has been connected to other interpersonal interaction indicators such as trust and patient satisfaction(7).Aestheticians introduced The concept of empathy in the mid-19th century by using the German word “Einfühlung”.Theodore Lipps,in1903, mean this concept as “feeling one’s way into the experience of another” Later, Martin Buber promoted this concept by describing empathic communication as “I and Thou,” instead of unempathic “I and It”(8) a struggle of words. Unfortunately, all of these definitions imply that empathic behavior is almost a revery etiquette.In medical settings, a wide spectrum of definitions and measures have covered empathy, however, there is no entirely accepted definition for empathy(9). Empathy can be explained as a multidimensional construct including ethical, affective, behavioral, and predominantly cognitive dimensions involving a patient’s understanding of experiences and concerns with an ability to communicate this understanding (rather than emotional feelings)(10). Several studies are defending empathy’s favorable health yields for clients.

The vulnerable situation of patients with hematologic malignancies, due to deterioration of their health, confronting the diagnosis, getting along with treatments and their side effects(11,12), quality of their life, and complex decision making in oncology settings, necessitate effective empathetic interaction in full attention from the physicians and who is working in oncology wards(13,14). Hematologic malignancies (HMs) are a miscellaneous group of malignant disorders that have crucial contributions to the cancer global burden(15). They are commonly distributed by their four main subtypes: leukemia, Hodgkin lymphoma (HL), non-Hodgkin lymphoma (NHL), and multiple myeloma (MM)(16). In 2018, leukemia accounted for 407,000 incident cases and 309,000 deaths(17) Moreover, in 2017, there were 1.4, 7.0, and 2.3 million disability-adjusted life-years (DALYs) for HL, NHL, and MM(14). this growing trend requires more intensive attention to these patients. Several clinical models have been introduced to integrate the EHR into patient care(18,19,20,21) however, recommendations are mostly based on anecdotal physician’s viewpoint rather than backed by the patient’s opinion in the highest level of evidence.

Based on recent statements, In our study, our main purpose was to compare patients’ perceptions of physicians’ empathy after the patient viewed standardized and scripted video vignettes of two physicians one uses the EHR system and the other one consults the patient without Examine Room Computer. Our second goal was to compare patients’ perception of patient-physician empathy and overall patients’ preference after watching each video and establishing the demographic and clinical predictors of patients’ preferences. Our guiding hypothesis was that patients would prefer physicians who don’t communicate with an EHR system and practicing with the EHR was reported as obtrusive and distracting (18,22,31). Eye contact as a fundamental nature of nonverbal social interaction has been studied in detail in past several studies(23,24,25,26,27,28,29). they also perceive physicians who visit the patient without Examine Room Computer, as more compassionate and having better communication skills(30). Therefore, this study was conducted to gauge the effect of Electronic Health Recording during visits on the level of physician-patient empathy in patients with hematological malignancies in February and March, 2023.

## Methods and materials

The study was registered and approved by the local Ethics Committee of Islamic azad university, Najafabad branch (IR.IAU.NAJAFABAD.REC.1401.170) and all participants signed informed consent. The current study was held in Omid hospital, Isfahan, Iran(Isfahan cancer center) aimed to assess the effects of EHR using on the physician-patient’s empathetic properties during visits using the Jefferson scale of empathy. In this study, after the patient viewed standard scripted video vignettes of physicians who portraying the use of an EHR(#1) and the other depicting consultation without using the examine room Computer(#2). The second goal was to compare patients’ perception of patient-physician empathy and overall patients’ preference after watching each video and establishing the demographic and clinical predictors of patients’ preferences.

### Patient Population

this is a double-blind, crossed-over clinical trial. patients were enlisted from the Hematology-oncology ward and Clinic of Omid hospital, Isfahan, Iran. (either new referrals or follow-ups).

### Samples

500 Patients were assessed for eligibility, 125 patients enrolled and 120 of them (96 percent) ultimately finalized the study. they were found to be eligible if they; spoke Farsi, were aged ≥18 years and had a diagnosis of early or advanced Hematologic cancer,(local, recurrent, or metastatic). the trained research staff evaluated patients’ cognitive status using The Memorial Delirium Assessment Scale (MDAS), patients with scores ≥7, and patients with severe physical/emotional impairment that were unable to attend the study were excluded(31).

### Blinding and Interventions

To minimize the biases, all patients were kept blinded to the study’s particular hypothesis, by asking them to observe 2 short videos and complete the questionnaire regarding the physicians acting in the videos. All coordinators and cast members were also blinded. The physician role was played by a man, aged 40 to 50 years. A 40 to 50-year-old woman with a poor prognosis, cancer played the patient role. The selection of the cast’s ethnicity and accent was determined by the familiarity of the Majority of patients referred to the Omid Hospital. The script represents routine discussions done in daily physician-patient encounters, which included evaluating the patient signs and symptoms, laboratory findings, medication prescriptions, and counseling. Each heading of the script had a specific communication skills empathic assessing statements with neutral emotions. The eye contact duration between the EHR-using section and the No Computer section was equivalent. The full script is available in the Supporting Materials. A biostatistician obtained the random assignment sequence.in the next step, Participants watched 2 video vignettes that were approximately 4 minutes long. In the first video, the physician is portraying the use of EHR during clinic visits; in the other video, the physician visits the patient without Computer. The recordings were observed and independently proved by six faculty members of the medical school of Isfahan University who were blinded to the main study’s hypothesis.

### Outcome measures

patient’s demographic and clinical traits were assessed then,the patients were asked to state the level of their agreement or disagreement with each of the Validated statements of the Persian translation of the Jefferson Scale of Patient Perceptions of Physician Empathy (JSPPPE) questionnaire that is recently developed for measuring patient perceptions of their physician’s empathy(36,37,50). The questionnaire consists of two parts, the first part includes personal and demographic characteristics and the second part is a five-point Likert-type scale (strongly agree:5, agree:4, have no opinion:3, disagree:2, strongly disagree:1). The scale consists of 24 items. total score ranges from 24 to 120; by Adding the points from the 24 statements together. The minimum possible score would be 60 and the maximum would be 120. the higher score indicates stronger empathy (33).

### A score between 24 and 40

the doctor’s level of empathy is low from the patients’ point of view.

### A score between 40 and 80

the doctor’s level of empathy is moderate from the patients’ point of view.

### A score higher than 80

the doctor’s level of empathy from the patient’s point of view is high. The reliability of this questionnaire has been measured by Managheb and Bagheri(36)and Cronbach’s alpha coefficient was estimated to be above 0.7. (This scale has high internal consistency, Cronbach α coefficient = .79)(37).

### Statistical analysis plan

Data were summarized using standard descriptive statistics, such as means, standard deviations and medians. Categorical variables were examined using the χ2 test or the Fisher test. Wilcoxon rank-sum test was used for continuous variables between groups. The generalized estimation equation (GEE) model was involved to assess the video effect and carry-over effect as well as demographic and clinical characteristic markers. The conception of the physician’s empathy was evaluated in a crossover study plan. All tests were 2-sided, and P ≤ .5 was assumed statistically significant. All data were entered into the SAS (Institute Inc) for computations.

## Results

120 patients were randomized into either the EHR or No-computer arms for a crossover study plan.

The first arm viewed the EHR(#1) and the second arm viewed No Computer (#2) (figure 1).

**Figure 1.**
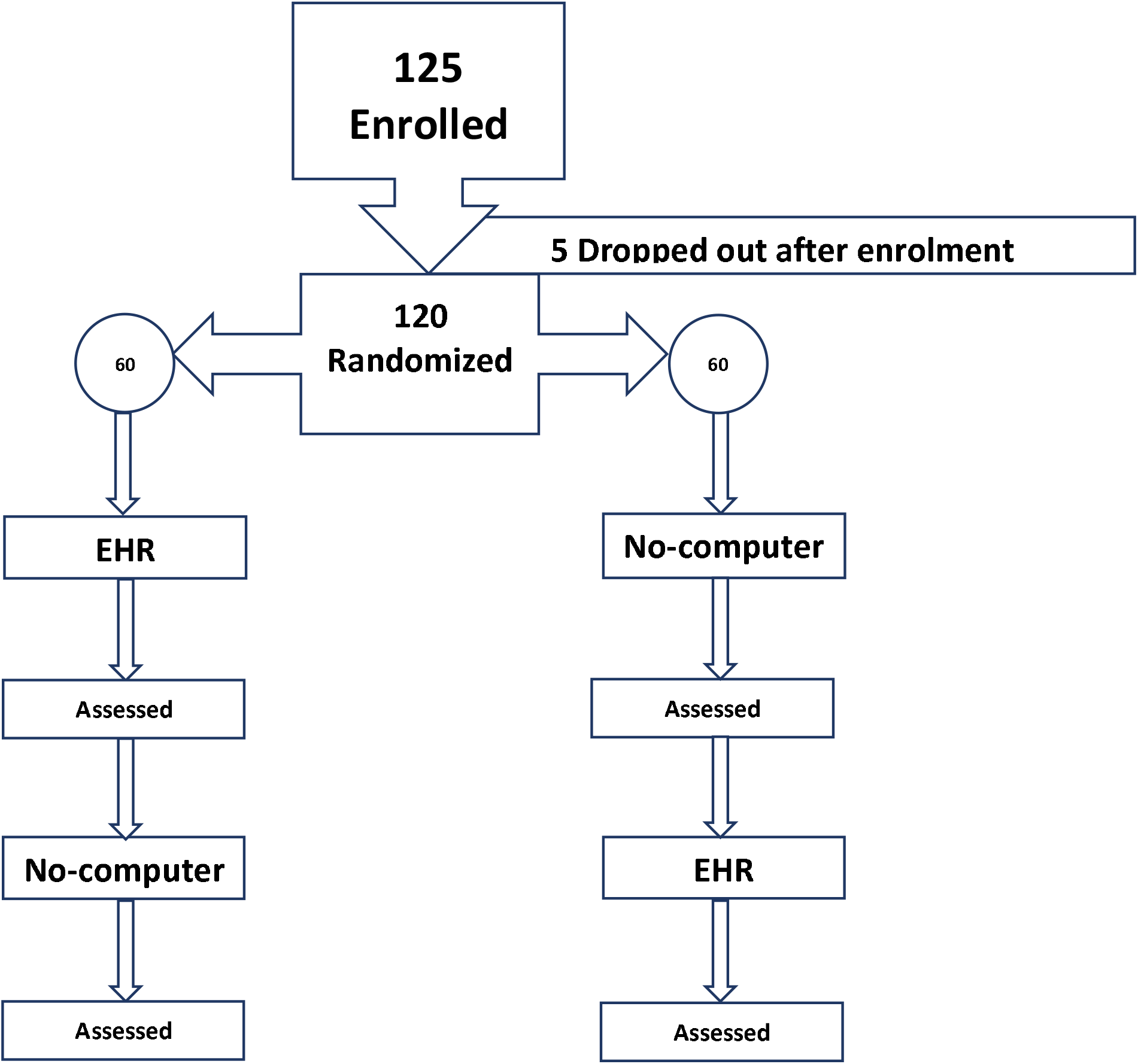
The study plan

Considering that the questionnaires were delivered to the patients by the research staff and completed there in their presence. The questionnaires were completed by all patients(51) and 100% of the them were returned for analysis. For illiterate patients, the questions were read by the research staff, and their answers were recorded.

Demographic characteristics of the patients are outlined in Table 1.

**Table 1.** Patients Demographic and Clinical Characteristics

| Variable | No. of Patients |
| --- | --- |
| Age: Median [IQR], y | 58 [27-74] |
| Sex |  |
| Women | 68(120) |
| Race |  |
| White. | 119(120) |
| Others | 1(120) |
| Marital status |  |
| Married | 98(120) |
| Unmarried | 22(120) |
| Education level |  |
| >Completed college | 76(120) |
| <College | 44(120) |
| Religion |  |
| Muslim | 114(120) |
| Other | 6(120) |
| Primary cancer diagnosis |  |
| Leukemia | 88(120) |
| Hodgkin Lymphoma | 4(120) |
| Non Hodgkin Lymphoma | 10(120) |
| Multiple Myeloma | 15(120) |
| Other. | 3(120) |
| Current stage |  |
| Metastatic | 14(120) |
| Recurrent | 30(120) |
| Locally advanced | 76(110) |

There were fewer men patients and more women patients,(men patients: 52[43%] vs women patients 68 [73%])، respectively; women show more empathy than men(53). There is a significant relationship between the grade of malignancy-prognosis and level of empathy percepted (p=.033).There were no further statistically strong differences in patient demographic characteristics and type of Primary cancer diagnosis with empathy scores.

## Main Outcome

After the two videos, the No Computer visit resulted in better empathy scores compared with the EHR visit (median score, 86; [interquartile range (IQR),67-105] vs 59 [IQR, 50-70];)P = .0009) (as featured in table 2).

**Table 2.** Outcome measures of physician empathy

| Outcome Measures: Patients Empathy Scores | No-computer visit | EHR visit | *P* |
| --- | --- | --- | --- |
| After first video :Median [IQR] | 80 [74-95] | 56 [49-64] | .0115 |
| After second video: Median [IQR] | 86 [67-105] | 59 [50-70] | .0009 |

Abbreviations: EHR, electronic health record; IQR, interquartile range.

The majority of patients (92 of 120; 77%) preferred the physician who didn’t use the computer for their care. in contrast, the Patients with advanced academic degrees (OR, 4.76; 95% CI, 1.34-17.04; P = .0172) were likely to favor the physician who uses the HER system.

## Discussion

The Empathy phenomena at its core will never change(48,49) but have various facets that are progressively being understood. Ultimately, it is crucial to understand how it influences their cognitive and physical outcomes. although it might be criticized from an ethical point of view that do good and avoid harm.

There are theoretical reasons to suspect both positive and negative effects of the Computer on doctor–patient interaction and relationships. The attention provided to patients with hematologic cancer in one-on-one encounters in the clinic is a fundamental part of their care(38). It can ultimately assist clinicians in supporting and guiding each individual patient throughout their cancer trajectory and establishes the trust between patients and physicians and strongly influences the success of long-term care through patients’ willingness to comply with prescribed treatments and continuous monitoring (39) these patients also see their clinicians as more empathetic (40).

As we continue to advance technological devices to foster patient care and outcomes, investigating and addressing obstacles to the expression of empathy is crucial(49). In this study, we highlight patients’ experiences and opinions regarding Iran as a predominantly religious-spiritual society(41) and the ubiquitous use of technology. physicians who communicated face to face without using the EHR were not only perceived as affirmative but were preferred by the patients.

Our findings suggest that by the No Computer approach, patients’ perceptions of their physician’s empathy were significantly fostered.

It elucidates that clinicians should decide on the best-suited approach for their clinical scenarios.

Certain No Computer visit aspects, such as proper introduction, better eye contact as the nature of communication skills besides digital modalities of EHR as an additional device (ie, images and laboratory tests to review, etc) may have had an affirmative impact on patients’ empathetic perceptions. it is possible that results would be different in patients due to their different computer literacy levels. In comparison with other skills, the skill of empathy will never be a simple one-size-fits-all recipe and depends to some extent on the personality of people(42,43,44). Some less significant factors related to the patient, such as age, gender, and race, can also influence the doctor’s empathetic responses to the patient’s negative emotions (39).

Ultimately, we continue to promote technological devices to improve patient care and outcomes. If medical care trends in The triumphs of technology, especially in hematooncological clinics resume as expected, empathy will become an even more critical issue.

thus exploring and addressing obstacles to the expression of empathy is crucial.

Further research is required to assess empathy fatigue in clinicians and the actual harmfulness of a lack of empathy in different clinical settings as digital modalities are advancing in different stages of cancer care (diagnosis and identification)(46,47).

A study in Philadelphia contrary to the Japanese one declares that the empathy of medical students recoils during clinical training (53). creating an empathetic relationship as a teachable skill (45, 46) may be forgotten in the curriculum of medical education, and gaining clinical experience without Proper training in these skills can have destructive effects on physician empathy and even make some physicians callous (48) thus further studies seem to be necessary on teaching these formulations systematically and continuously (43).

Words themselves do not contain wisdom. Words said to particular individuals at particular times may occasion wisdom.

—Iris Murdoch

## Data Availability

All data produced in the present study are available upon reasonable request to the authors.

https://ethics.research.ac.ir/ProposalCertificateEn.php?id=314305&Print=true&NoPrintHeader=true&NoPrintFooter=true&NoPrintPageBorder=true&LetterPrint=true

## Declaration

One of the major practical obstacles of this research was the concern of the patients about not receiving proper services from the doctor if they refused to fill out the form correctly, which was explained with sufficient explanation and assurance that all the information in the questionnaire is protected and their participation in this study is voluntary.

Their worries would be solved and they would do the proper cooperation.

The authors would like to thank MS the faculty member of Isfahan university of medical sciences and all other faculty members and patients who participated in this study. The study was registered and approved by the local Ethics Committee of Islamic azad university, Najafabad branch (IR.IAU.NAJAFABAD.REC.1401.170) and all participants gave their informed consent.

Authors certify that have no affiliations with or involvement in any organization or entity with any financial interest or non-financial interest in the subject matter or materials discussed in this manuscript.

Conceptualization, investigation, methodology, project administration, resources, supervision, validation, visualization, writing-original draft, and writing-review and editing were done by SADT.

SADT:read and approved the final manuscript.

Funding:None.

Raw data were generated at Isfahan University of medical sciences. Video vignettes, questionnaires and derived data supporting the findings of this study are available from the corresponding author [SA] on request.

